# *SERPINA11* related novel Serpinopathy- a perinatal lethal disorder

**DOI:** 10.1101/2023.10.03.23296272

**Authors:** Shagun Aggarwal, Venugopal Satidevi Vineeth, Shrutika S. Padwal, Sameer Ahmed Bhat, Arpita Singh, Aditya Kulkarni, Mallikarjun Patil, Karthik Tallapaka, Divya Pasumarthi, Vijayasree Venkatapuram, Ashwin Dalal, Rashna Bhandari

## Abstract

*SERPINA11* is a hitherto poorly characterised gene belonging to Clade A of the SERPIN superfamily. The exact functional significance and expression pattern of this gene remains unknown. Here we report a perinatal lethal phenotype associated with biallelic loss of function variants in *SERPINA11*, and characterised by gross and histopathological features of extracellular matrix disruption. We found SERPINA11 protein expression in multiple mouse tissues and human fetal lungs. Immunofluorescence of the affected fetal lung revealed markedly reduced expression of SERPINA11 protein compared with healthy, gestation matched human fetal lung. Protein expression data from HEK293T cell lines following site directed mutagenesis is also presented to support the loss of function nature of the variant. This novel serpinopathy appears to be a consequence of loss of inhibition of serine proteases involved in extracellular matrix remodelling, revealing SERPINA11 as a protease inhibitor critical for embryonic development.

## Main text

The conserved SERine Protease INhibitors (SERPIN) superfamily is composed of 37 genes divided across nine (A-I) clades.^1-5^ Some of these proteins have predominant inhibitory roles in regulating various serine proteases in the extracellular matrix, while others play important roles in coagulation, inflammation and immune response (Inhibitory SERPINs), and some others act as binding molecules, hormone precursors and chaperones (Non-inhibitory SERPINs). All these proteins share a common tertiary structure although the degree of sequence homology is highly variable.^1-5^ Alpha 1 antitrypsin deficiency (MIM: 613490) due to biallelic pathogenic variations in *SERPINA1*(MIM:107400) is the most common and well characterised serpinopathy.^6; 7^ This condition presents with a phenotype of pulmonary emphysema and liver cirrhosis. The pathophysiology involves uninhibited activity of neutrophil elastase (a serine protease) resulting in destruction of lung tissue, and PAS positive aggregates of the abnormally folded SERPINA1 in liver cells resulting in hepatic damage.^6-10^ Other delineated serpinopathies are Thrombophilia due to antithrombin III deficiency (MIM:613118) caused by pathogenic variants in *SERPINC1*(MIM:107300); Angioedema, Hereditary, type I and II (MIM:106100) caused by pathogenic variants in *SERPING1*(MIM:606860); and Encephalopathy, familial, with neuroserpin inclusion bodies (MIM:604218) caused by pathogenic variants in *SERPINI1*(602445). This is the report of a novel serpinopathy due to biallelic loss of function variants in Serpin Peptidase Inhibitor, Clade A, Member 11 (*SERPINA11*), a previously poorly categorised member of this family.

A non-consanguineous couple presented in 20-25th week of pregnancy with ultrasound done elsewhere showing isolated pericardial effusion (Figure 1A). There was history of a previous term neonate with intractable congenital pleural effusion who succumbed in 0-2 months of life. In view of this neonatal outcome, the uncertain prognosis of the ultrasound finding was discussed with the couple and amniocentesis was performed. The couple terminated the pregnancy and the fetus (affected fetus 1; f1) was brought for a post-mortem evaluation. On external examination, there was mild subcutaneous edema with subtle facial dysmorphic features like puffy eyes, horizontal crease at the nasal root, infra orbital creases, broad nose, long philtrum and mild micrognathia. Thickening of alae nasi and ear lobules was appreciated. Right foot was inverted and splaying of toes was noted. The neck, chest, abdomen and spine were normal. The external genitalia were normal male. The placenta appeared normal with a three vessel cord. Anthropometric measurements corresponded to 50-95th centile of 24 weeks. Intra thoracic dissection revealed minimal serous pericardial effusion. A small gelatinous glistening cyst was present on the right pericardium. Bilateral pleural effusion was noted. Multiple similar cysts were noted on the lung surfaces as well, more in the dependent portions. Intra-thoracic structures were structurally normal otherwise. Intra-abdominal structures were grossly normal but for similar thin walled cystic lesions containing honey coloured gelatinous fluid noted over surface of the liver and along the intestines. No specific diagnosis could be made on the basis of these findings (Figure 1B-1E).

**Figure 1:**
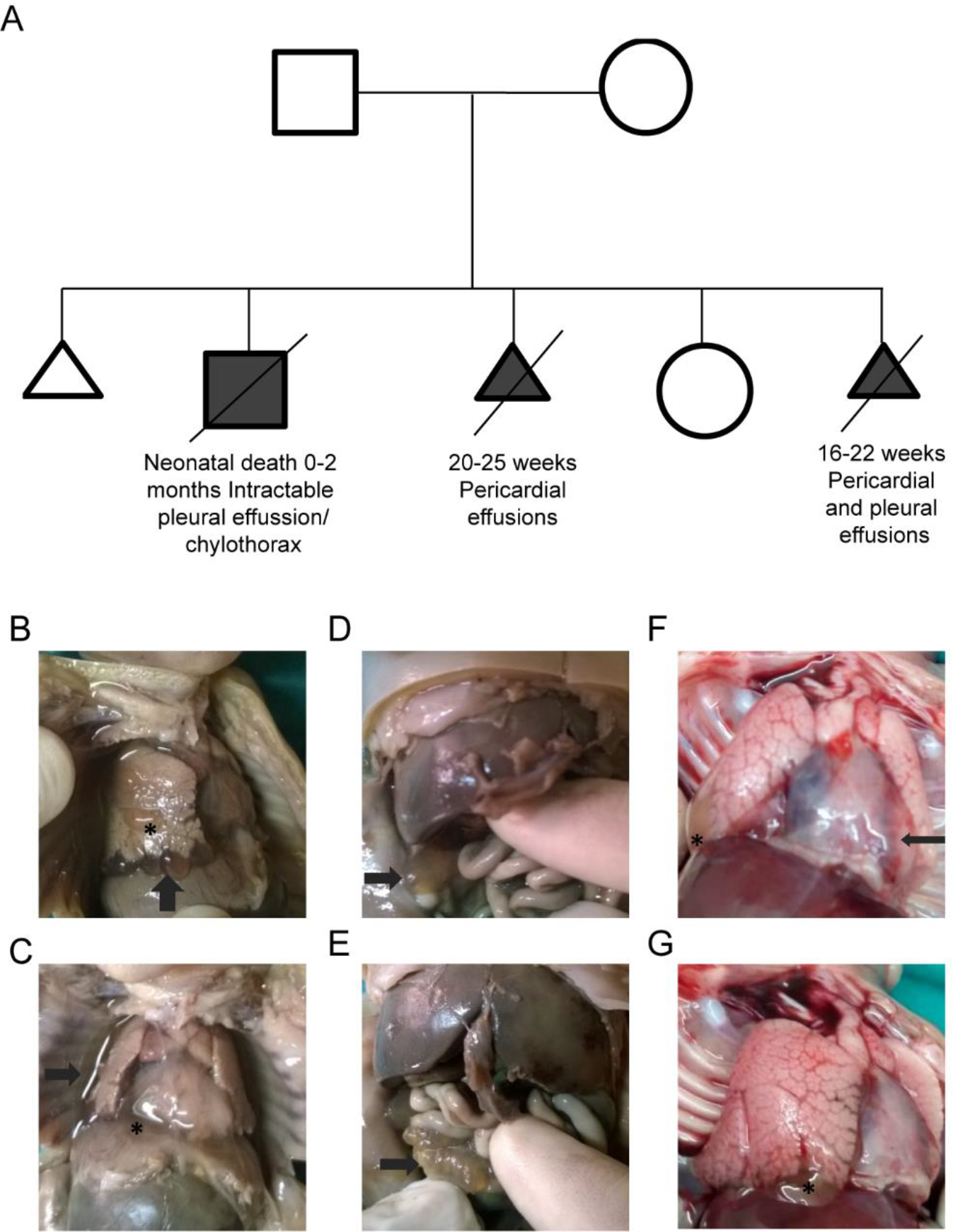
Autopsy photographs of the affected fetuses and family pedigree. (A) Pedigree of the family. (B) Right lung of fetus 1 showing presence of multiple blebs lined by thin membrane on the anterior surface (asterisk) and under-surface (arrow). (C) Presence of thin membrane lined cyst/bleb in pericardium (asterisk) and right lateral pleural surface (arrow). (D) Presence of similar thin membrane lined cyst/bleb on liver under-surface (arrow). (E) Presence of multiple thin membrane lined blebs on mesentery (arrow). (F) Presence of thin membrane lined cyst/bleb in pericardium (arrow) and right lateral pleural surface (asterisk) in fetus 2. (G) Right lung of fetus 2 showing presence of similar blebs lined by thin membrane on the under-surface (asterisk).

The study was approved by the Nizam’s Institute of Medical Sciences Institutional Ethics Committee (Review Letter Numbers EC/NIMS/1555(a)/2015 and EC/NIMS/2232/2018) and all human samples were obtained after informed consent. MLPA for common aneuploidies (13, 18, 21, X and Y) in fetus and parental karyotypes were normal. Solo whole exome sequencing was performed from stored amniocyte DNA. DNA isolated from blood (∼1 μg) was used to perform exome capture using the Nextera Rapid Capture Exome v1.2 Kit (Illumina, San Diego, CA) following the manufacturer’s protocol. The libraries were sequenced to >100× coverage on Illumina HiSeq2000 platform (Illumina, San Diego, CA). Exome sequencing and data analysis pipelines were described earlier.^11^ Human genome build hg37 was used for mapping and annotation. Targeted PCR was performed using gene-specific primers [Forward Primer 5’ GCCAAGGCATTTCCTCTGTA 3’; Reverse Primer 5’ TGGCCTTATGGGTTTTGTGT3’] and Sanger sequencing was done using ABI 3130 Genetic analyzer (Life Technologies, Carlsbad, CA). Pathogenicity of the variant was tested using prediction tools like MutationTaster2 and CADD.^12; 13^

A homozygous nonsense variant in exon 3 of a candidate gene Serpin Family A Member 11 *(SERPINA11)*, NM_001080451.2;c.672C>A;p.Tyr224*, was shortlisted as likely causative of the fetal phenotype (Figure S1A). This variant was absent from population databases like 1000 Genomes, gnomAD and our in-house database. The variant was predicted pathogenic by in-silico prediction tools like SIFT, Polyphen and Mutation taster, with a CADD score of 38.^14; 15^ The quality information of exome sequencing raw data and the variant filtering strategy are outlined in Supplemental Table S1 and S2 respectively. The gene has not been linked to any known disorder in humans as per OMIM. Sanger sequencing revealed both the parents to be carriers for the same variant (Figures S1B, C).

*SERPINA11* was hypothesized to be a candidate gene on basis of it belonging to the clade A of SERPIN superfamily, similar to *SERPINA1. SERPINA1* or alpha1 antitrypsin deficiency involves a lung phenotype of emphysema which is characterised by the destruction of walls of the terminal airways due to uninhibited neutrophil elastase activity.^6; 7^ The phenotype of the fetus was somewhat reminiscent of a similar extracellular matrix abnormality with prominent involvement of lung surfaces. In addition, another research group had reported a biallelic splice variant as VOUS in *SERPINA11* in a fetus with hydrops, although detailed post-mortem phenotype of this case was not available.^16^ We further performed histopathological evaluation of the fetal organs using Haematoxylin-Eosin and Masson Trichrome stains, to look for evidence of extracellular matrix involvement (Figure 2A-H). Sections from bilateral lungs showed predominantly canalicular phase of lung maturity, corresponding to the gestational age. The pleural space appeared cystically enlarged and showed focal formation of bullae with expansion of the pleural stroma. These areas showed disruption of collagen fibers and the background matrix. These foci also showed breakdown of elastin. Similar expansion and disruption were also seen in peribronchiolar areas. The liver similarly showed occasional bullae on the capsular surface, with expansion of the capsular stroma and breakdown of collagen and other components of the ECM. The stroma around the central vein and portal triads also showed similar findings. The hepatic parenchyma showed normal hepatocyte arrangement with sinusoidal erythropoiesis. Both the kidneys showed normal cortico-medullary differentiation with a well – defined outermost nephrogenic zone. The peripelvic connective tissue showed expanded matrix with loss of normal architecture/framework. All other fetal organs were unremarkable. No PAS positive inclusions were appreciated in the hepatocytes.

**Figure 2:**
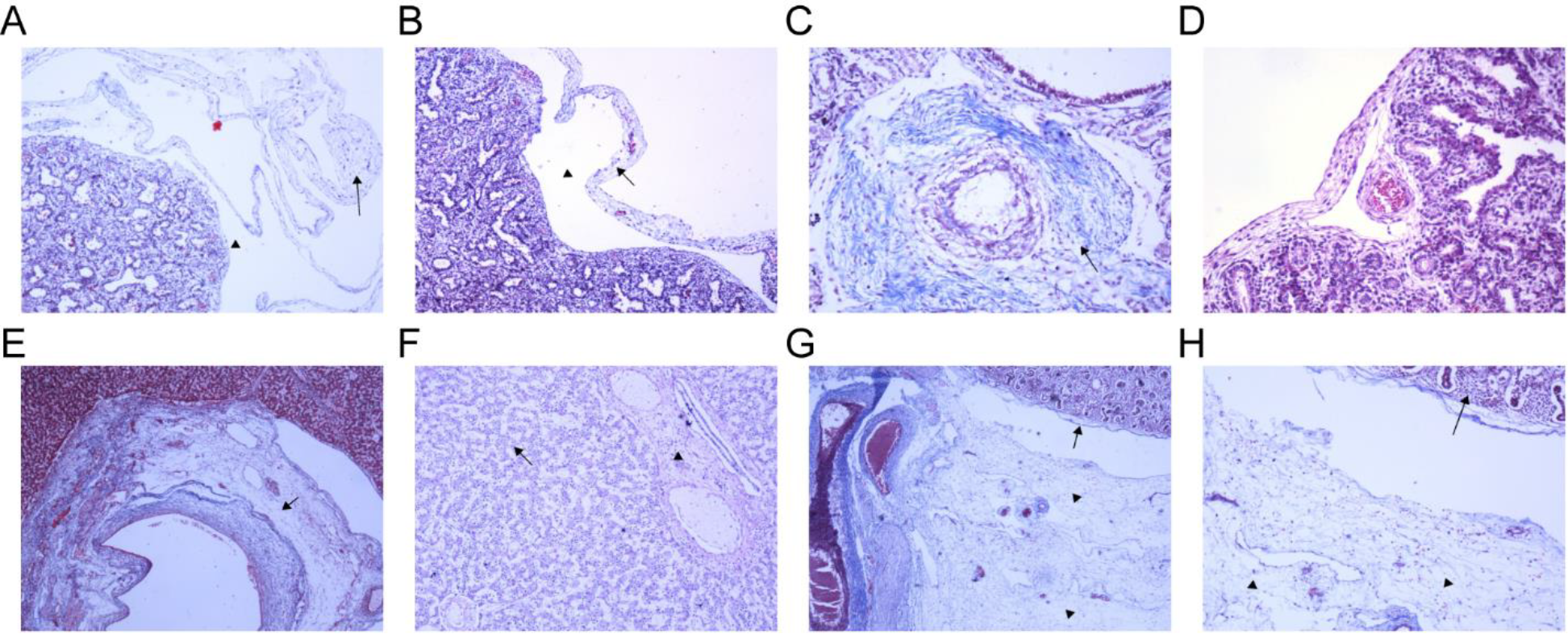
Histological staining of lung, liver and kidney sections. Histological staining of gestational age matched affected and normal fetuses. (A, B) Affected fetus 1 lung section showing blebbing / bulla formation on the pleural surface (arrowhead). The pleural stroma shows expansion with fragments of collagen (arrow); Masson’s trichrome staining, 4x (A), 10x (B). (C) Affected fetus 1 peribronchiolar connective tissue shows expanded matrix and loss of normal collagen architecture (arrow); Masson’s trichrome staining, 20x. (D) Normal fetal lung pleura with no blebbing for comparison; Hematoxylin-Eosin staining, 20x. (E) Affected fetus 1 liver section showing hepatic hilum with expansion of the supporting matrix and disruption of normal architecture of collagen (arrow); Masson’s trichrome staining, 4x. (F) Normal fetal hepatic parenchyma for comparison with normal collagen surrounding the central vein (arrowhead) and hepatocytic trabeculae (arrow); Hematoxylin-Eosin staining, 4x. (G, H) Affected fetus 1 kidney section showing expansion of peripelvic connective tissue with fragmentation of collagen (arrowheads). Renal parenchyma showing developing nephrons is seen in the upper corner (arrow); Masson’s trichrome staining, 4x (G), 10x (H).

These findings indicated significant disruption of the extra cellular matrix supporting the candidature of *SERPINA11* as a Mendelian gene responsible for the phenotype. We hypothesize that this gene product may act as an inhibitory SERPIN involved in regulating serine proteases and is responsible for extracellular matrix homeostasis. A literature search revealed that information regarding the tissue expression, biological functions and structure of the corresponding protein was minimal. The predicted protein has 422 amino acid residues with 41% amino acid sequence homology with SERPINA1 (Alpha1 antitrypsin) and a predicted molecular weight of ∼47kDa.^17^ A 19 residue N-terminal signal peptide is predicted to target SERPINA11 to the endoplasmic reticulum for subsequent trafficking to the extracellular matrix. SERPINA11 also possesses four N-linked glycosylation sites. 3D structure prediction using AlphaFold^18; 19^ (https://alphafold.ebi.ac.uk/entry/A0A4W2C737) suggests that SERPINA11 adopts a similar tertiary structure as SERPINA1^20^ with an exposed reactive centre loop (RCL) (Figure S2). The RCL plays a vital role in protease inactivation by acting as a pseudo-substrate for the target protease.^4^ Cleavage of the scissile bond between P1 and P1’ residues in the RCL by the target protease leads to a conformational change in the serpin. Following this, the serpin either remains covalently bound to the protease and inactivates it, or is itself inactivated and releases the active protease. The ratio between the two alternative pathways is expressed as the stoichiometry of inhibition and should be close to 1 for SERPINs to become powerful inhibitors.^1-5^ The RCL of SERPINA11 is predicted to be formed by residues 366-397 (ebi.ac.uk/interpro/protein/UniProt/Q86U17/). The structure of the c.672C>A;pTyr224* SERPINA11 variant predicted using ColabFold^21^, shows that this variant lacks the RCL domain (Figure S2), and hence is predicted to cause a complete loss of function.

Lysates from HEK293T cells transiently over expressing C-terminally eGFP-tagged SERPINA11 were subjected to western blotting and probed with an anti-GFP antibody or anti-SERPINA11 antibody (Supplemental materials and methods). Expression of eGFP-tagged SERPINA11 at ∼75 kDa was detected by both antibodies (Figure 3A-B), corresponding accurately to the mass of GFP (∼27 kDa) fused with SERPINA11, which has a predicted molecular weight of ∼47kDa based on its primary sequence. Site directed mutagenesis of *SERPINA11* cDNA was conducted to recreate the identified variant by replacing the codon for Tyr224 with a stop codon. Western blot analysis of lysates from HEK293T cells transfected with this Tyr224* mutant construct did not detect truncated SERPINA11, suggesting that the epitope recognised by this commercially available anti-SERPINA11 antibody lies C-terminal to Tyr224* (Figure 3A-B). An antibody to GFP also failed to detect any protein expression from this Tyr224* construct, indicating that the stop codon is functional. However, on flow cytometry analysis, HEK293T cells transiently over expressing C-terminally eGFP-tagged SERPINA11 Tyr224* showed 0.1% GFP positive cells, indicating partial read through of the variant which was not detected by western blotting (Figure 3C). There was no change in transcript levels of mutant over wild type *SERPINA11* in transfected HEK293T cells, confirming that loss of expression of the Tyr224* mutant is not due to reduced transcription (Figure 3D).

**Figure 3:**
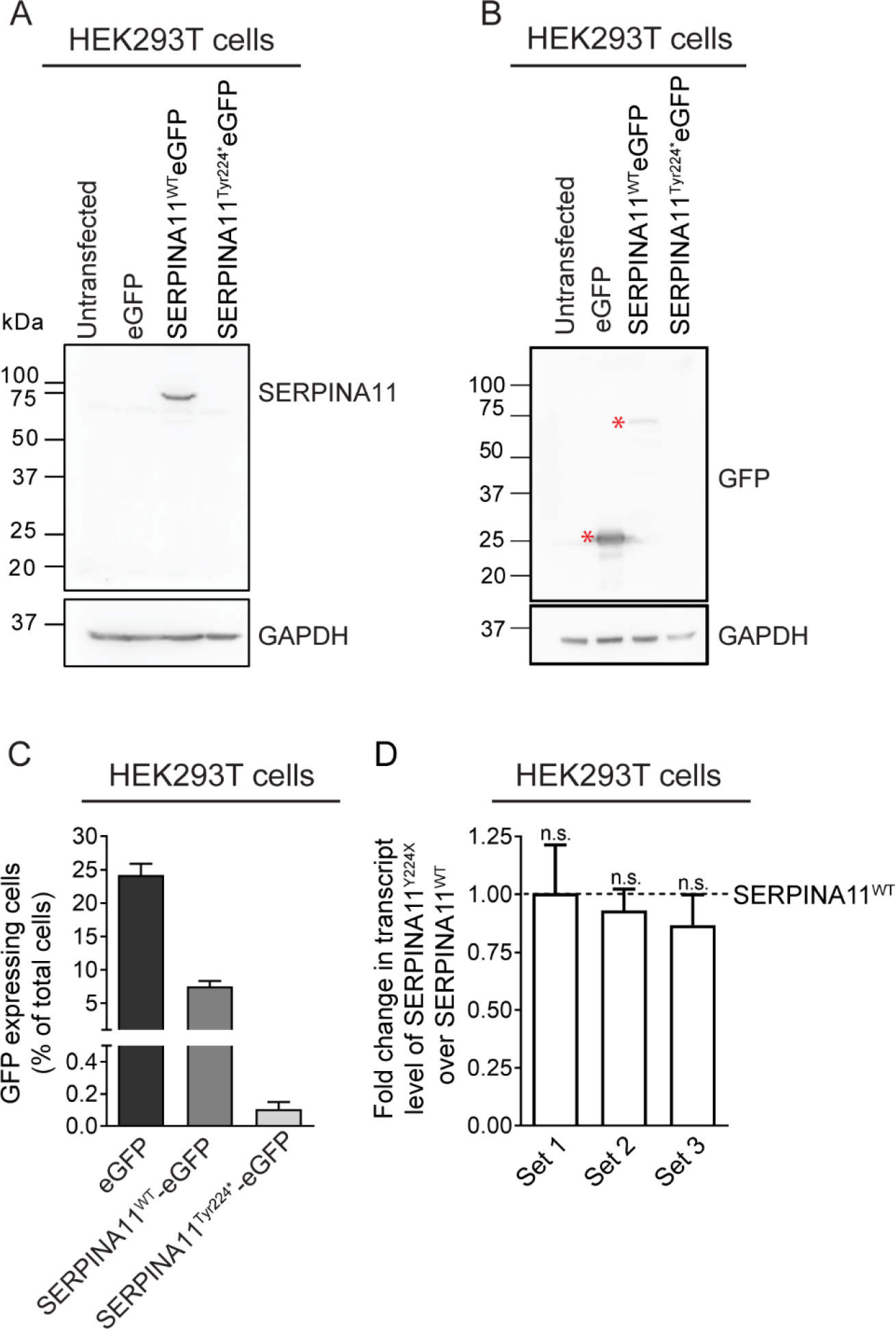
Functional validation of SERPINA11 Tyr224* nonsense variant. (A-B) Representative immunoblots examining the expression of SERPINA11 in HEK293T cells transiently overexpressing wild type and Tyr224* mutant SERPINA11, C-terminally tagged with eGFP, and probed with anti-SERPINA11 antibody (A) or anti-GFP antibody (B). Correct size bands were observed for eGFP and wild type SERPINA11-eGFP. An asterisk (*) indicates the specific band. GAPDH was used as the loading control. (C) Flow-cytometry analysis of HEK293T cell lines transiently expressing the same proteins as in (A, B), showing percentage of GFP expressing cells. (D) RT-qPCR analysis, using three different primer sets, of *SERPINA11* transcripts in HEK293T cells transfected with plasmids encoding wild type and Tyr224* mutant SERPINA11. Data are mean±s.e.m (N=4). P-values were determined using a one-sample t-test. P≤0.05 was considered significant, n.s., not significant (P>0.05).

We conducted western blot analysis to examine expression of SERPINA11 in liver, lung, kidney, heart, brain, testis, and ovary of adult C57BL/6 mice (Figure 4A). A∼47kDa protein corresponding to the predicted molecular weight of mouse SERPINA11 was detected in mouse lung, liver, and testis, whereas single bands of higher molecular weight were detected in mouse brain, heart and kidney. The apparent shift in molecular weight is suggestive of differential glycosylation of SERPINA11 in these tissues. The expression pattern of SERPINA11 in mice mirrors the tissues in which gross pathologies were observed in the affected fetus. Although *Serpina11* transcript has been reported only in the liver of adult C57BL/6 mice (https://www.ebi.ac.uk/gxa/), our detection of the protein in different mouse tissues could arise from SERPINA11 transport to these tissues via circulation, or low level of *Serpina11* transcription in these tissues.

**Figure 4:**
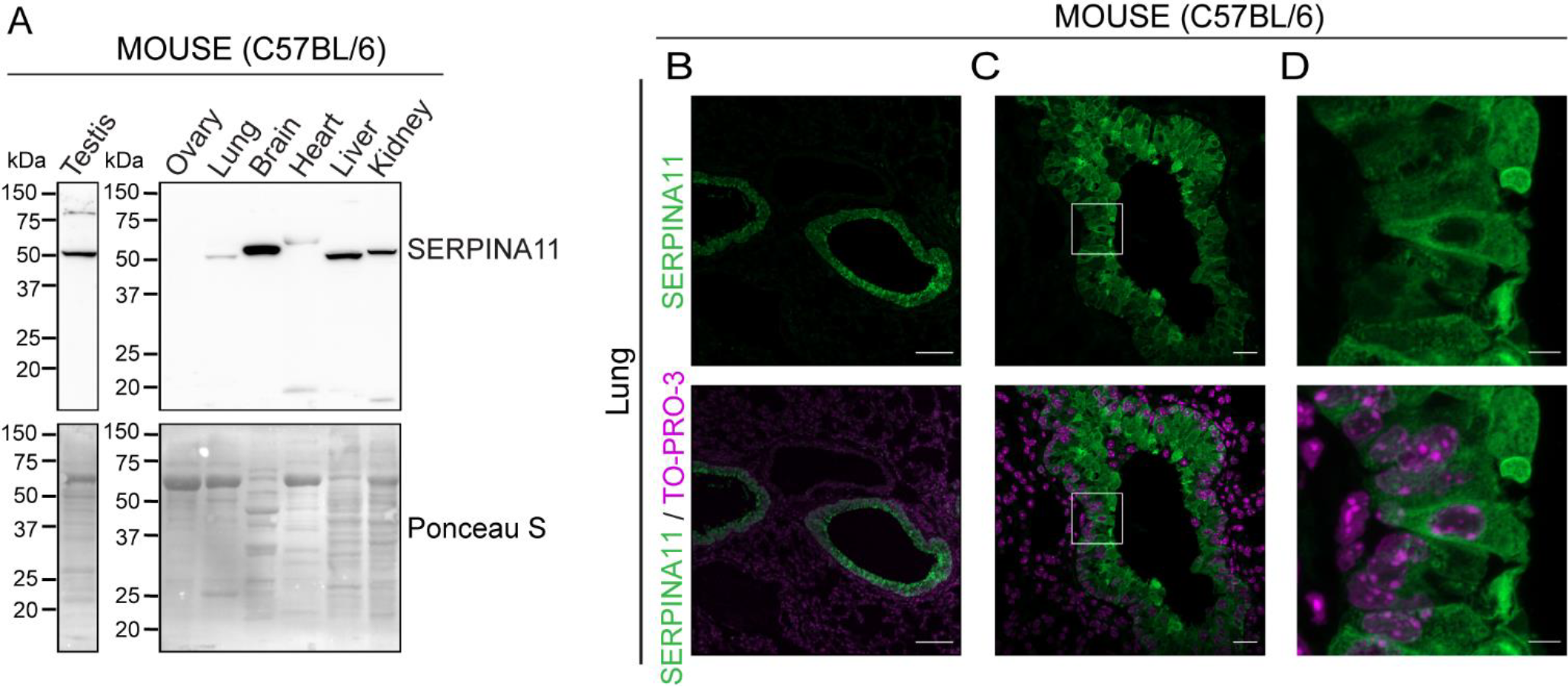
SERPINA11 is expressed in multiple mouse tissues. (A) Representative immunoblots examining the expression of SERPINA11 in lung, brain, heart, liver, kidney, ovary, and testis of adult male and female mice. SERPINA11 was detected using anti-SERPINA11 antibody. Total protein detected by Ponceau S dye was used as the loading control. (B-D) Representative immunofluorescence images of lung sections at different magnifications from two month old adult male mice showing SERPINA11 (green). Nuclei were detected with TO-PRO-3 stain (magenta). The boxed area in (C) is shown in (D). SERPINA11 staining is seen in the cytoplasm of bronchiolar pseudostratified columnar epithelial cells and is absent in alveolar epithelia. Scale bar 100μm (B), 20μm (C), 5μm (D).

To identify the cell types expressing SERPINA11 we performed immunofluorescence analysis in lung, kidney and liver sections of adult mice (Supplemental materials and methods). Staining of these tissue sections with the same anti-SERPINA11 antibody used for western blotting, revealed an intense signal in the pseudostratified bronchiolar epithelium in mouse lung (Figure 4B-D). Interestingly, we observed intracellular staining of SERPINA11 in bronchiolar epithelial cells, which is suggestive of *Serpina11* transcription within these cells. The alveolar epithelia did not show any SERPINA11 staining. We were unable to detect an immunofluorescence signal in mouse liver and kidney (Supplemental Figure S3), despite robust expression of SERPINA11 detected by western blotting in these tissues. This could be due to diffuse localization of the protein in liver and kidney.

Finally, we conducted immunofluorescence analysis of lung sections from the affected fetus and a gestational age matched normal fetus. SERPINA11 expression was clearly detected in the bronchiolar epithelium in the normal fetus, but was undetectable in the affected fetus (Figure 5).

**Figure 5:**
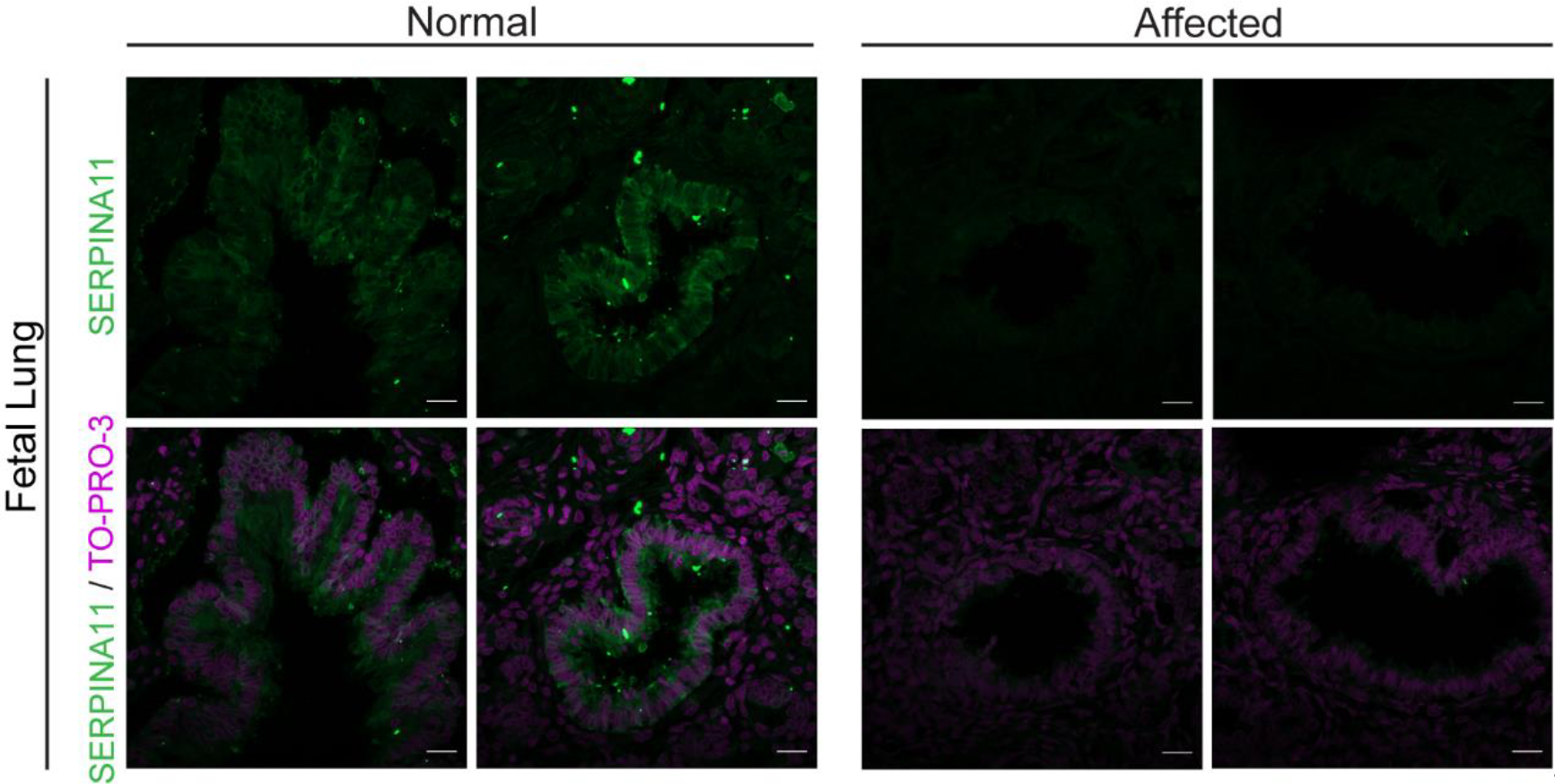
SERPINA11 is undetectable in affected fetal lung tissue. Representative immunofluorescence images of lung sections of affected fetus 1 and gestational age matched normal fetus were stained to detect SERPINA11 (green), and nuclei were detected with TO-PRO-3 stain (magenta). Scale bar 20μm.

During the course of the work, the couple conceived twice, one pregnancy resulted in birth of a healthy female child, while in the subsequent pregnancy, ultrasonography at 16-22 weeks gestation showed pleural and pericardial effusion (Figure 1A). This pregnancy was terminated and female fetus (affected fetus 2; f2) was brought for postmortem evaluation. Gross findings of f2 were similar to f1 (Figure 1 F, G). Sanger sequencing on fetal DNA revealed a homozygous state for the c.672C>A variant (Figure S1D). In the case of f2, we were able to retrieve unfixed tissue samples, suitable for RNA extraction and analysis. We compared *SERPINA11* transcript levels in fetal liver, lung and kidney from f2 with the human hepatocellular carcinoma cell line HepG2, which has been reported to express *SERPINA11* (https://www.proteinatlas.org). Interestingly, *SERPINA11* mRNA was detectable in all three tissues from f2 (Figure S4).

Our findings indicate that *SERPINA11* is a fully functional gene that is expressed in various tissues in the developing human fetus and adult mice. Based on sequence similarity and predicted structure, SERPINA11 appears to belong to the class of inhibitory serpins, although its target proteases are undefined at present. The phenotype presented here is a perinatal lethal one. The nature of the variant, confirmation of protein truncation by site-directed mutagenesis, and fetal lung immunofluorescence data suggest that the lethal phenotype results from a complete loss of function of the protein. Histologically, there is generalised marked extracellular matrix disruption seen in various internal viscera connective tissue and visceral membranes. Hence, SERPINA11 appears to be involved in extracellular matrix homeostasis by inhibitory action on various tissue serine proteases. Also, the protein appears to be ubiquitous and plays an important role during embryonic development, unlike other serpins, many of which have roles primarily in the postnatal period, and disruptions of which cause relatively tissue specific phenotypes^1-8^. Further studies are needed to define the target proteases of SERPINA11 and to determine whether it is primarily an intracellular or secretory protein. Structural biology approaches are needed to identify the SERPINA11 structure, characterize its functional domains and their interactions with proteases. Finally, as serpinopathies are being targeted for treatment using recombinant serpin therapies and other molecular approaches, the feasibility of such an approach as an in-utero therapy for SERPINA11 related phenotype may also be envisaged in future.^10^

To conclude, we report a novel serpinopathy which is a perinatal lethal phenotype of extracellular matrix disruption resulting from a biallelic loss of function variant in *SERPINA11* gene.

## Data availability

The *SERPINA11* variant was submitted to ClinVar (https://www.ncbi.nlm.nih.gov/clinvar/) and is available for viewing via accession number VCV000631492.3. The exome sequencing dataset supporting the study has not been deposited in a public repository but are available upon request to the corresponding author.

## Author contributions

Conceptualization, S.A., A.D., V.S.V., and R.B.; Methodology, S.A., V.S.V., S.S.P., S.A.B., A.S., and R.B.; Validation, S.A., V.S.V., S.S.P., S.A.B., A.S., and R.B..; Formal analysis, S.S.P., A.S., and R.B.; Investigation, S.A., V.S.V., S.S.P., S.A.B., A.S., A.K., K.T., M.P., D.P. and V.V.; Writing original draft preparation, S.A., V.S.V., S.A.B., and R.B.; Writing review and editing, S.A., S.S.P., and R.B.; Visualization, S.A., V.S.V., S.S.P, S.A.B., A.S., and R.B; Supervision, S.A., A.D, and R.B; Project administration, S.A., A.D, and R.B; Funding acquisition, S.A. All authors have read and agreed to the published version of the manuscript.

## Acknowledgments

The authors thank the family for their kind consent and for providing samples for the research study. We acknowledge funding support from the Science and Engineering Research Board (SERB), Government of India (Grant/Award Number: EMR/2016/002478), Department of Health Research (DHR), Government of India (Grant/Award Number: File R.12020/05/2020-HR/E-Office:8055238), and CDFD core funds. S.S.P. and A.S. are recipients of Junior and Senior Research Fellowships from the Council of Scientific and Industrial Research, and Department of Biotechnology, Government of India, respectively.

## Web resources

SIFT, https://sift.bii.a-star.edu.sg/

PolyPhen2, http://genetics.bwh.harvard.edu/pph2/

Mutation Taster, http://www.mutationtaster.org/

CADD, https://cadd.gs.washington.edu/

gnomAD, https://gnomad.broadinstitute.org/

OMIM, https://omim.org

EMBOSS Needle, https://ebi.ac.uk/tools/psa/emboss_needle/

AlphaFold, https://alphafold.ebi.ac.uk/

ColabFold https://github.com/sokrypton/ColabFold

InterPro, https://www.ebi.ac.uk/interpro/

Expression atlas, https://www.ebi.ac.uk/gxa/home

Human Protein Atlas, https://www.proteinatlas.org

