## Supplemental Methods, Tables, and Figures for "*SERPINA11* related novel Serpinopathy- a perinatal lethal disorder"

### SUPPLEMENTAL INFORMATION

#### Supplemental Materials and Methods

**Reagents:** All chemicals were procured from Sigma-Aldrich, unless specified otherwise. The primary antibodies used in this study for immunofluorescence (IF) and immunoblotting (IB) along with antibody dilution for each application and supplier (including catalogue number) are as follows: anti-SERPINA11 (Invitrogen, PA5-50755; IF 1:100; IB 1:1000), anti-GAPDH (Sigma-Aldrich, G8795; IB 1:10,000), anti-α-tubulin (Sigma-Aldrich, T9026; IB 1:10,000), anti-GFP (Invitrogen, A11122; IB 1:5000). Restriction enzymes were purchased from New England Biolabs. Dulbecco's Modified Eagle's Medium (DMEM), Fetal Bovine Serum (FBS) and other cell culture reagents were from Thermo Fisher Scientific.

**Plasmids:** The human *SERPINA11* ORF clone procured from GenScript (OHu55737D) was sub-cloned into the Xho1 and EcoR1 restriction enzyme sites in the pEGFPN1 expression plasmid to encode C- terminally GFP tagged SERPINA11. Site directed mutagenesis to generate the c.672C>A mutation was conducted using Phusion high fidelity DNA polymerase (Thermofisher) with mutagenic oligonucleotides **[**Forward Primer 5' CTGCTTCTGGGTCTGTTAGCGACTGAAAGGGT 3'; Reverse Primer5' ACCCTTTCAGTCGCTAACAGACCCAGAAGCAG 3'**]**. PCR for the substitution were run for 24 cycles of 95°C for 45s, 68°C for 30s and 72°C for 5min. The resulting mutant plasmids were verified by sequencing.

**Cell lines and transfection:** All cell lines were grown in a humidified incubator with 5% $CO_2$ at 37°C in Dulbecco's Modified Eagle's Medium (DMEM) supplemented with 10% fetal bovine serum, 1 mM L-glutamine, and 100 U/mL penicillin and 100 µg/mL streptomycin. Wild type and mutant human SERPINA11 encoding plasmids were transfected into HEK293T cell line using polyethylenimine (PEI) (Polysciences, 23966) at a ratio of 1:3 (DNA:PEI). All plasmids used for transfection were purified using the Plasmid Midi kit (Qiagen). Cells were harvested 48h post-transfection for further analyses.

**Mice:** All animal experiments were approved by the Institutional Animal Ethics Committee (Protocol number EAF/RB/CDFD/25), and were performed in compliance with guidelines provided by the Committee for Control and Supervision of Experiments on Animals, Government of India. Mice (*Mus musculus*, strain C57BL/6) used for this study were housed in the Experimental Animal Facility at the Centre for DNA Fingerprinting and Diagnostics, Hyderabad. Adult 12 week old male and female mice were euthanised by carbon dioxide inhalation followed by collection of brain, heart, lung, liver, kidney, ovary and testis.

**Western blot analysis:** Transfected HEK293T cells were collected after 48 h by briefly washing with phosphate buffered saline (PBS) and mixed with 4X SDS sample buffer to a final concentration of 1X. Mice organs were dissected, briefly washed in chilled PBS, and minced in RIPA buffer (50 mM Tris-HCl pH 7.4, 150 mM NaCl, 1% Triton X-100, 0.5% sodium deoxycholate, 0.1% SDS, 1 mM EDTA, 10 mM NaF, 1 mM PMSF) containing freshly added protease inhibitor cocktail (Sigma-Aldrich). The tissues were subjected to homogenisation (D-1 Homogenizer-Disperser, Miccra) and sonication (Bioruptor, Diagenode). The extracts were centrifuged at 20,000 *g* for 15 min at 4°C and supernatants were immediately mixed with 4X SDS sample buffer to a final dilution of 1X. The samples were heated at 95°C for 5 min, resolved by SDS-PAGE and transferred onto PVDF membranes (Cytiva). Western blotting was performed to detect the protein of interest and to assess equal gel loading by sequential probing of blots with experimental and control antibodies. In the case of mouse tissues, equal gel loading was confirmed by staining the blots with Ponceau S prior to blocking, as the expression of standard loading control proteins (e.g. actin, tubulin, GAPDH) may vary between different tissues. Membranes were blocked with 5% skimmed milk for 1 h at room temperature, followed by incubation with the primary antibody overnight at 4°C. After washing three times with TBST buffer (20 mM Tris-HCl pH 7.6, 150 mM NaCl and 0.1% Tween 20), membranes were incubated at room temperature for 1 h with HRP-conjugated secondary antibodies as follows: goat anti-rabbit IgG conjugated to HRP (Southern Biotech, 4010-05; 1:5000) for SERPINA11 and GFP; goat anti-mouse IgG conjugated to

HRP (Southern Biotech, 1030-05; 1:5000) for GAPDH and α-tubulin. After washing the membrane three times with TBST, specific proteins bands were detected using a chemiluminescence reagent (ECL Prime, Cytiva). Chemiluminescence was detected using the ImageQuant LAS 500 documentation system. Where required, blots were stripped using RestoreTM Western Blot Stripping Buffer (ThermoFisher Scientific), and re-probed with the control antibody. All western blotting experiments were conducted for two to three biological replicates.

**Immunohistochemistry:** After isolation, all mouse and human fetal organs were briefly washed with PBS and fixed in 10% neutral buffered formalin (NBF) solution for 24 h at room temperature (RT). Representative sections were then sampled from these tissues and dehydrated in increasing concentrations of alcohol, followed by clearing with xylene and finally infiltration with paraffin wax. The processed sections were then embedded in paraffin blocks. 4 µm sections were cut from the blocks using a microtome (Leica RM2125 RT) and taken on a glass slide. The paraffin-embedded sections were deparaffinised in a hot air oven (65°C), cleared in xylene, and rehydrated with decreasing concentrations of alcohol before undergoing antigen retrieval by boiling in 10 mM sodium citrate buffer (pH 6) in a microwave oven for 20 min. The sections were then washed in PBS, permeabilised in 0.5% Triton X-100 for 10 min, blocked in blocking buffer (4% FBS+0.1% Triton X-100) for 1 h at RT and then incubated overnight at 4°C with anti-SERPINA11 antibody diluted (1:100) in blocking buffer. The slides were washed with PBS and stained with Alexa Fluor 488-conjugated goat anti-rabbit-IgG (Molecular Probes, 1:500 dilution in blocking buffer), for 1 h at RT. Nuclei were stained using TO-PRO-3 (ThermoFisher, T3605) at 1:1000 dilution for 15 min. The slides were washed with PBS and mounted in antifade mounting medium (H 1000, Vector Labs). Images were acquired with an LSM 900 confocal microscope (Zeiss, Zen acquisition software) equipped with 405, 488, 561, and 633 nm lasers, and fitted with a 63X 1.4 NA objective. Parallel sections from the tissue blocks were stained with Hematoxylin and Eosin using standard protocols.

**RT-qPCR:** HEK293T cells were transiently transfected with plasmids encoding either SERPINA11 WT or SERPINA11 Tyr224*. Total cellular RNA was isolated from the transfected cells using TRIzol reagent (Invitrogen) followed by RNeasy Mini Kit (Qiagen). 2 µg of total RNA was used for first strand cDNA synthesis by reverse transcription using SuperScript Reverse transcriptase III (Invitrogen) with oligonucleotide dT primers. qPCR was performed using 3 sets of *SERPINA11* primers (Set 1, Set 2, Set 3) and *GAPDH* specific primers on the ABI 7500 Real-Time PCR System (Applied Biosystems) with DyNAmo ColorFlash SYBR Green qPCR Mastermix (ThermoScientific) for detection. Each sample was run in triplicate. The difference in transcript levels between mutant and wild type *SERPINA11* was calculated using the fold change ($\Delta\Delta C_t$ method). $\Delta C_t$ is the $C_t$ value for

*SERPINA11* WT or Tyr224* normalized to the $C_t$ value of the respective *GAPDH* control. $\Delta\Delta C_t$ values were calculated as a relative change in $\Delta C_t$ of *SERPINA11* Tyr224* with respect to *SERPINA11* WT. Fold changes were expressed as $2^{-\Delta\Delta Ct}$.

To quantify *SERPINA11* RNA in fetal tissues, cDNA was prepared from total RNA isolated from lung, liver, and kidney from f2, and from HepG2 cells as a positive control. qPCR was conducted using Set 3 *SERPINA11* primers. $\Delta\Delta C_t$ values were calculated as a relative change in $\Delta C_t$ of *SERPINA11* in RNA from each tissue with respect to RNA from HepG2 cells.

Primer sequences:

*SERPINA11*:

Set 1: Forward primer (FP) 5’-TGTTACAACTGGGAGGCAGA-3’, Reverse primer (RP) 5’-CCACTTGGCTTTGAAGAAGATG-3’; Set 2: FP 5’-ACACGTTCATGGTTCTTGCC-3’, RP 5’-TTTTGGTGCATCATGGGGAC-3’; Set 3: FP 5’-CAGTCTGTTGGATTTGCACTTG-3’, RP 5’-TTTTGTTGAGCTGCCCAGTG-3’

*GAPDH*:

FP 5’-ACATCAAGAGCTGGTGAAGC-3’, RP 5’-TTGTCATACCAGGAAATGAGCTTG-3’

### Supplemental Tables

**Table S1.** Quality information of exome sequencing raw data.

| | |
|---|---|
| Total number of reads | 70876693 |
| Total reads mapped (%) | 70663211 (99.7%) |
| Reads that passed alignment (%) | 70370318 (99.29%) |
| Mean depth of coverage[1] | 140X |
| Quality threshold[2] | 90.9% |

[1]Mean depth of coverage refers to the sequence mean read depth across the targeted region, defined as coding exons and splice junctions of CaptureReagent kit targeted protein coding RefSeq genes.
[2]The quality threshold refers to the percentage of the defined target region where read depth was at least 10X coverage.

**Table S2.** Filtering strategy for variants observed in whole-exome sequencing.

| **Filtering criteria** | **Number of variants** |
|---|---|
| Total number of variants | 102184 |
| Variants remaining after filtering for polymorphisms | 8794 |
| Variants remaining after filtering for synonymous variants | 8279 |
| Variants remaining after filtering for non-coding regions variants with lack of evidence | 930 |
| Variants remaining after filtering out low priority variants (pathogenicity prediction tools) | 611 |
| Variants in known genes that can explain the phenotype and recurrence | 0 |
| Number of variants in homozygous state | 35 |
| Homozygous variants having CADD score ≥ 10 | 17 |
| Homozygous variants in genes with previous report in abnormal fetus but not functionally characterized | 1 |

### Supplemental Figures

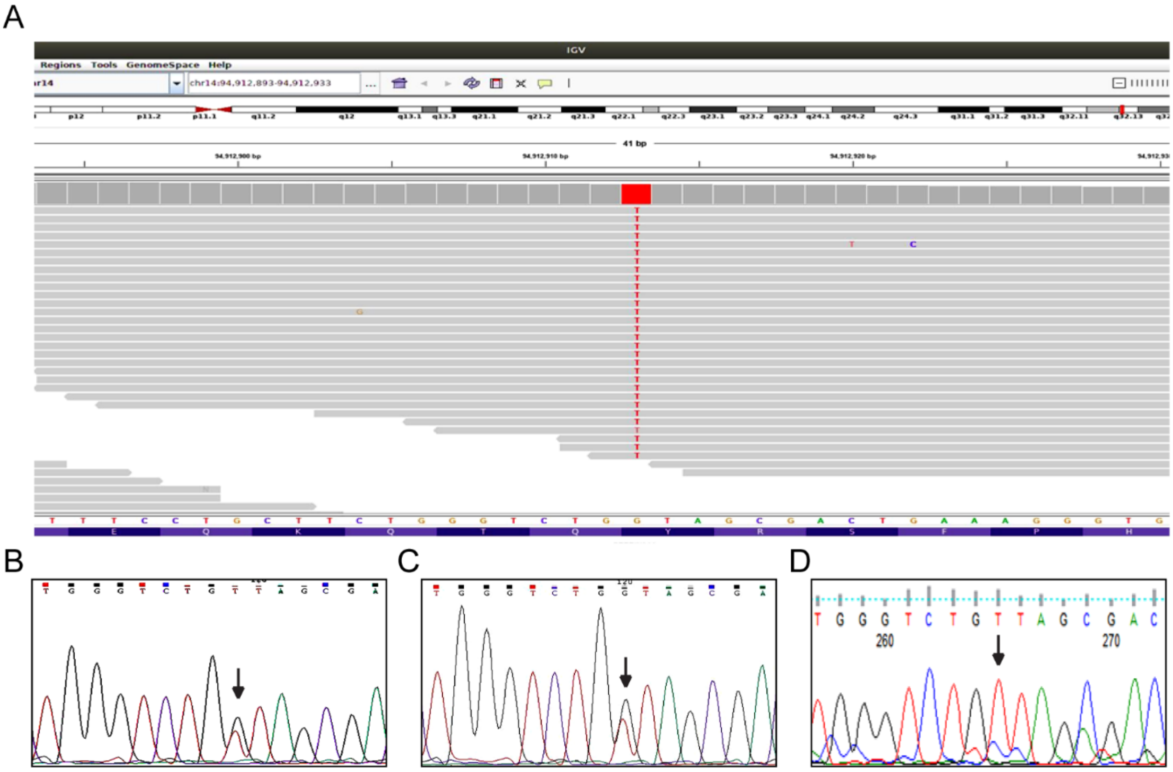


**Supplemental Figure 1: *SERPINA11* sequence analysis** (A) Figure showing Integrative Genomics Viewer (IGV) screenshot of the *SERPINA11* variant (NM_001080451.2;c.672C>A;p.Tyr224*) in fetus 1. (B-D) Sanger sequencing showing heterozygous status (NM_001080451.2;**c.672C>A**) in father (B) and mother (C), and homozygous status (NM_001080451.2;**c.672C>A**) in fetus 2 (D).

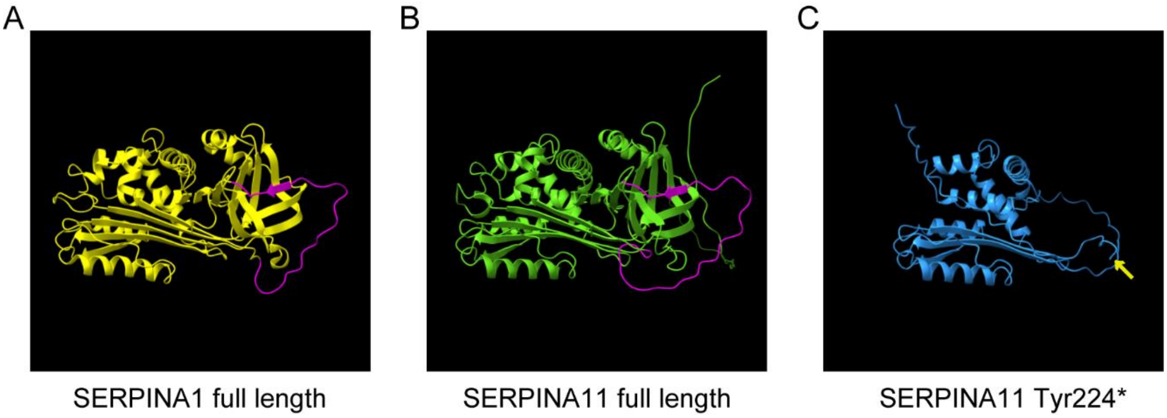


**Supplemental Figure 2: SERPINA11 structure prediction.** (A) Tertiary structure of wild type human SERPINA1 residues 43-418 (PDB ID: 1HP7). (B) Structure of full length human SERPINA11 predicted by Alphafold2. (C) Predicted structure of Tyr224* mutant SERPINA11 obtained using ColabFold. The reactive centre loop (RCL) in (A, B) is marked in magenta. The arrow in (C) indicates the truncation in Tyr224* mutant SERPINA11. Structures were visualized using the UCSF Chimera tool.

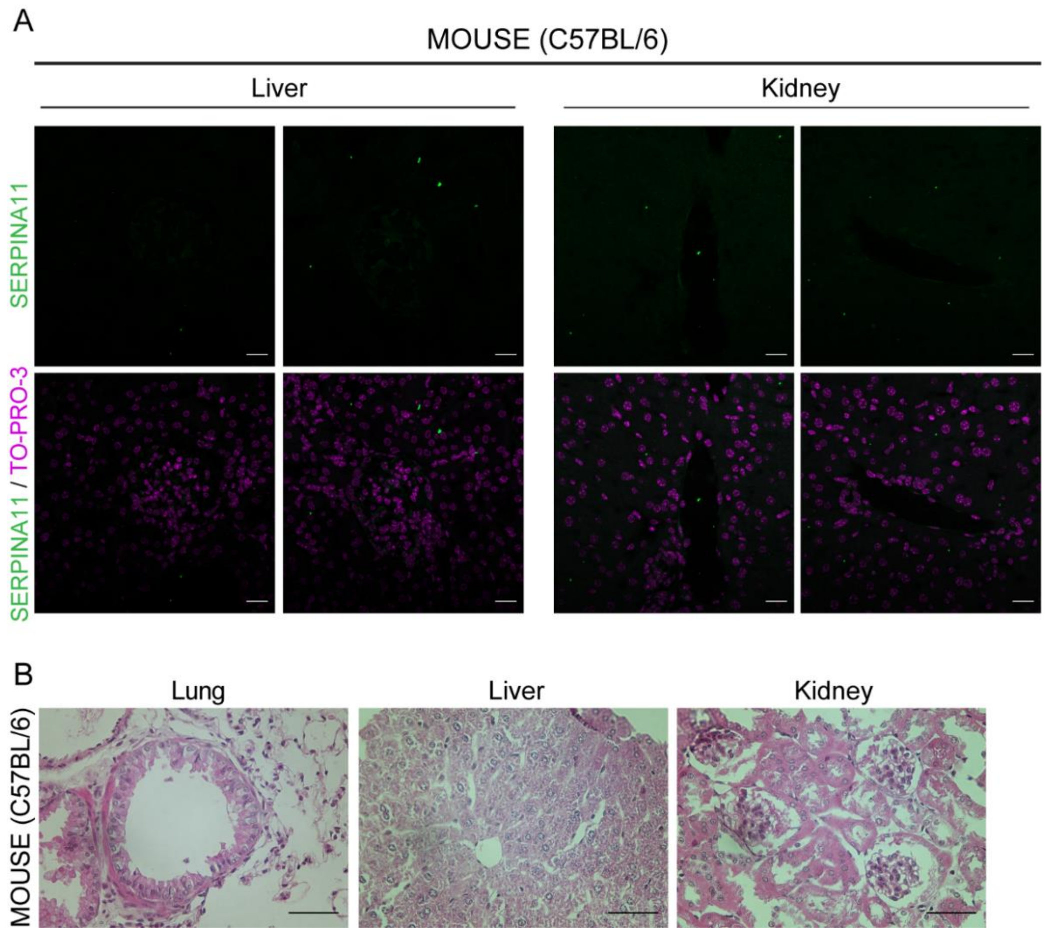


**Supplemental Figure 3: SERPINA11 staining is not detectable in mouse liver and kidney.** (A) Representative liver and kidney sections from two month old adult male mice were stained to detect SERPINA11 (green). Nuclei were detected with TO-PRO-3 stain (magenta). (B) Hematoxylin and Eosin staining of mouse tissue sections adjacent to those stained in (A) and Figure 4B-D, showing normal tissue architecture. Scale bar 20µm (A), 50 µm (B).

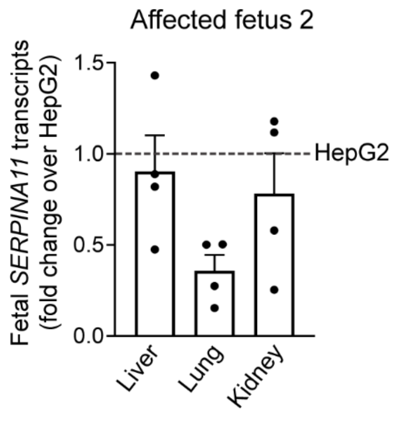


**Supplemental Figure 4: Fetal *SERPINA11* RNA analysis.** RT-qPCR analysis of *SERPINA11* transcripts in RNA from liver, lung and kidney tissues from affected fetus 2, compared with HepG2 cells.
